# Housing instability among young men who have sex with men in a US national sample

**DOI:** 10.1101/2024.05.09.24307116

**Authors:** Amanda Sisselman-Borgia, Viraj V Patel, Christian Grov

**Affiliations:** CUNY Lehman College Department of Social Work; Division of General Internal Medicine, Albert Einstein College of Medicine, Montefiore Health System; Graduate School of Public Health and Health Policy, City University of New York, 55 W 125th St, 7th Floor mailroom., New York, NY, 10027 United States

**Keywords:** STI prevention, sexually transmitted infections, adolescents, youth, housing instability, homelessness, trauma

## Abstract

**Background:** Youth, including those experiencing housing instability, are among the fastest growing groups of individuals with new STI diagnoses, including HIV. The unpredictable nature of the lives of youth experiencing housing instability often leads to inconsistent or non-existent health care and preventive follow up, leaving gaps in our knowledge about the most prominent needs for intervention.

**Methods:** Using data from the *Together 5000* (T5K) study, we examined factors associated with housing instability in a sample of 2,228 youth between the ages of 16-24 who identified as sexual and gender minority (SGM) men having sex with men. Logistic regression was used to assess the most prominent factors associated with housing instability. The model included seven significant factors - former peer victimization, gender, age, sex work, IPV, social support, and health insurance status.

**Findings:** Participants who reported more behavioral risk factors for STI and those who reported sex work in the last three months were more likely to experience housing instability (OR = 2.5 and 2.76 respectively). Youth with higher levels of reported social support, health insurance, and older age were more likely to report stable housing (OR = .98, 1.61, and 1.13 respectively). Surprisingly, youth with stable housing were more likely to report intimate partner violence than those experiencing housing instability (OR = .89).

**Discussion:** Implications for addressing STIs among adolescent SGM men having sex with men are discussed including bolstering support systems and addressing basic needs deficits and trauma associated with sex work and behavioral risk factors for STI.

**Implications and contributions statement:** The study examines correlates of housing instability among a sample of young sexual and gender minority men who have sex with other men. Correlates of housing instability included behavioral risk factors for sexually transmitted infection and reporting sex work in the last three months. Health insurance, older age, and higher levels of social support were correlated with more stable housing.

## Introduction

Youth homelessness and housing insecurity is growing in the United States - 4.2 million children and young adults are impacted annually by physical, mental, and emotional impacts of being unhoused ^1^. Within New York City, between 2015 and 2018 there was a 16% increase in youth experiencing homelessness bringing the nightly shelter count of youth to 10,000^2^, a number that is likely a substantial underestimate given that many youth sleep in public spaces, such as subways or on the street each night ^3^. Youth who identify as LGBTQ+ (Lesbian, gay, bisexual, transgender, Queer, Intersex, Asexual) are also at a higher risk for becoming homeless and contracting HIV than heterosexual youth^4^. LGBTQ+ youth experiencing homelessness or housing insecurity also have high rates of trading sex for food, housing, or other necessities leading to additional victimization on the streets ^4^. For example, 75% of transgender youth reported periods of homelessness; and are more likely to engage in survival sex with multiple partners and injection drug use (IDU), increasing risk of HIV infection^5,6^.

LGBTQ+ youth experiencing housing instability are at a higher risk for health and mental health problems including post-traumatic stress and are heavily impacted by health disparities and racism^1,7,8^. Behavioral risk factors such as sex work and injection drug use, are heavily associated with homelessness especially among trans gender youth, putting youth experiencing housing instability at an increased risk for sexually transmitted infections (STIs)^6,9–11^. Youth, including those who are experiencing homelessness or housing insecurity, are among the fastest growing groups of individuals with new STI diagnoses, including HIV^12,13^. The unpredictable nature of the lives of youth experiencing housing instability often leads to inconsistent or non-existent health care and preventive follow up. Unhoused youth are less likely to have access to condoms and STI testing and may engage in condomless sex, which puts them at a higher risk for STI^14^. Relatedly, LGBTQ+ youth experiencing housing instability often engage in sex work or sex trade in order to barter for food, housing, and other basic needs. Experiences related to homelessness or housing instability including sex work, community violence, and sexual assault are also associated with physical health issues such as STIs and additional trauma and post-traumatic stress^15^.

Although there have been numerous studies conducted examining correlates of housing instability and homelessness among youth experiencing housing instability overall and LGBTQ+ youth who experience housing instability grouped together, there have been no national studies to our knowledge, focusing specifically on youth experiencing housing instability who identify as sexual and gender minority (SGM) men who have sex with men. The current study seeks to help fill this gap by examining the factors associated with housing instability among a large national cohort of young SGM men who have sex with men experiencing housing instability in order to be able to identify the public health problems needing most attention in this vulnerable population. In this paper, we focus on SGM men having sex with men to learn more about correlates of housing instability in this specific population, to control for confounding variables and identify possible factors that are unique to SGM men having sex with men that might get masked in a more general sample. We examine potential factors, including social support, associated with housing insecurity.

## Methods

### Participants and Procedures

The data used in this analysis came from the *Together 5000* (T5K) study, which used an internet-based survey to collect data from a U.S. national cohort focused on men having sex with men (MSM) who were HIV vulnerable. Cisgender MSM, trans men, and trans women who have sex with men were also included. Enrollment and research protocols are described elsewhere^16,17^. In 2017-2018, participants were recruited through ads on geosocial networking apps used by MSM. Participants were deemed eligible if they were between the ages of 16 and 49, reported at least two male sex partners in the last 90 days, not taking PrEP, self-reported negative HIV status, and report behavioral risk for HIV (i.e., engaged in receptive condomless anal sex or insertive condomless anal sex or engaged in injection drug use). A total of 8,674 people were eligible to enroll in the study of which 2,228 completed the baseline survey and were between the ages of 16 and 24 years old. All study procedures were approved by the T5K study PI’s Institutional Review Board.

## Measures

### Main outcome variable

Housing instability, our main outcome variable, was assessed through one question, “Has there been a period of time in your life when you have been unstably housed (including couch surfing)?” Response options ranged from “yes, within the last 5 years”, “yes more than 5 years ago”, and “no”. For the purposes of this analysis, we created a dichotomous housing instability variable, where “yes” indicated any history of housing instability. We hypothesized that housing instability would be associated with increased intimate partner violence (IPV), increased behavioral risk factors for STIs, decreased social support, increased former peer victimization (i.e., bullying), decreased resilience, and decreased LGBT+ support systems.

### Hypothesized factors Associated with housing insecurity

STI history was measured by asking participants if they had been diagnosed with a sexually transmitted infection in their lifetimes (yes/no). History of sex work was assessed by asking participants if they have exchanged sex for money or other items in the last three months (yes/no). Health insurance status was assessed with three options (yes/no/I don’t know). Behavioral risk for sexually transmitted infection was measured using six items developed by the T5K study team,^18^ which asked about engagement in behaviors such as injection drug use and condomless sex within the last 12 months. Participants responded yes or no and the items were summed to create a total risk index (range 0 to 6).

Resilience was measured using the Connor-Davidson Resilience Scale^19^, 10-item version. Responses are rated on a 5-point likert scale, ranging from 0 (not true at all) to 5 (true nearly all the time) and responses are summed, with a total range 0-50. Examples of items are, “I am able to adapt to change” and “I can handle unpleasant feelings” (chronbach alpha = .87). Use of LGBT+ resources was measured using seven items ascertaining how frequently participants visited LGBT+ friendly places, such as houses of worship, coffee shops, and healthcare providers (Cronbach alpha = .69); responses were measured on a 5-point likert scale, ranging from 0 (never) to 4 (more than 10 times), with a range of 0-28. Examples of items were, “In the last 6 months, how often did you visit a nearby LGBT community center?; In the last 6 months, how often did you visit a nearby LGBT+ friendly house of worship?”. Social support was measured using the Multidimensional Scale of Perceived Social Support (Cronbach alpha = .94)^20^. There were a total of 12 items, asking participants to rate their agreement on a 6-point Likert scale, ranging from 1 (very strongly disagree) to 6 (very strongly agree) with a range of 1-72.

Intimate partner violence (IPV) was measured using an adaptation of the Revised Conflict Tactics Scale (Cronbach alpha = .91)^21 22^. Participants were asked if they experienced any of a range of forms of IPV within the last five years. Twelve items were answered yes or no, and a total score was formed by summing the items, with total range of 0-12. Example items were, “hit with an open fist” and “verbally threatened you in any way”. Former peer victimization was measured using 12 items from the Multidimensional Peer Victimization Scale (Cronbach alpha = .91)^23^. Participants were asked to think back to when they were in high school and respond to how often a peer did certain actions to them on a three-point scale, with responses 0 (not at all), 1 (once), and 2 (more than once) with total range 0-24. Example items were, “called me names”, “refused to talk to me”, and “beat me up”.

#### Sample

For this analysis, we included only participants aged 16 through 24 years old, including those who identified as transgender (n = 2,228).

#### Analysis

We used descriptive statistics to report the demographic characteristics of the sample and to examine distributions and ensure statistical assumptions were met as necessary. Due to small sample sizes, we included gender in the multi-variable analysis as a dichotomous variable (cisgender male vs. not). We then conducted bivariate analysis using independent *t*-tests or chi-squared tests with our primary outcome variable (housing instability vs not) and hypothesized associated factors (included former peer victimization, resilience, LGBT+ resource use, health insurance status, IPV score, behavioral risk for sexually transmitted infection, sex work in the past 3 months, STI history, and social support). We then created a multivariable logistic regression model by including all factors hypothesized to be associated with housing instability. A binomial logistic regression was conducted to assess the impact of a number of factors on the likelihood that young SGM men having sex with men (cis gender, transgender male and female) between the ages of 16 and 24 would report housing instability. Our main hypothesis was that housing instability among young SGM men having sex with men (including Cis gender and transgender men and women) would be associated with increased intimate partner violence (IPV), increased behavioral risk factors for STIs, including sex work or exchange in the last 3 months, decreased social support, increased former peer victimization (i.e., bullying), decreased resilience, and decreased LGBT+ support systems.

## Results

### Sample Characteristics

Table 1 reports demographic data. A total of 2,228 (26% of the overall T5K sample) youth and young adults between the ages of 16 and 24 were included in our analysis, 569 (31%) of whom reported a history of housing instability. Our sample was primarily non-white among youth experiencing housing instability and those not experiencing housing instability (64% non-white and 60% non-white respectively). The majority of the sample had at least a high school diploma in both groups and most participants were either working full or part time.

**Table 1.** Demographic differences between those having experienced housing instability versus not.

| Variable | Experienced housing instability (n = 569 )<br>n (%) | Stably housed (n = 1648)<br>n (%) | $\chi^2$ |
| --- | --- | --- | --- |
| Gender |  |  | 41.36* |
| Male | 520 (91%) | 1604 (97%) |  |
| Transfemale | 17 (3%) | 8 (.5%) |  |
| Transmale | 8 (1.4%) | 9 (.5%) |  |
| Something else | 24 (4.2%) | 27 (2%) |  |
| Race |  |  | 12.28** |
| White | 203 (36%) | 662 (40%) |  |
| Black | 125 (22%) | 258 (16%) |  |
| Latina/o | 164 (29%) | 493 (30%) |  |
| Multi-racial | 77 (14%) | 235 (14%) |  |
| Education |  |  | 71.87** |
| Less than High School | 43 (8%) | 83 (5%) |  |
| High School or Equivalent | 163 (29%) | 348 (21%) |  |
| Some College | 184 (32%) | 391 (24%) |  |
| Enrolled in College | 125 (22%) | 539 (33%) |  |
| College degree or higher | 54 (9.5%) | 287 (17.2%) |  |
| Employment |  |  | 90.13** |
| Full time | 213 (37%) | 641 (39%) |  |
| Part-time (including FT students) | 201 (35%) | 630 (39%) |  |
| Disabled | 9 (1.5%) | 7 (.5%) |  |
| Unemployed | 146 (25%) | 370 (33%) |  |
| Health Insurance (yes response) | 233 (59%) | 857 (74%) | 33.10** |
| Sex work in past 3 mos (yes response) | 149 (36%) | 179 (15%) | 86.93** |
| Lifetime STI (yes response) | 219 (38.5%) | 515 (31.3%) | 10.01* |
| Age | M = 22 SD = 2.00 | M = 21 SD = 2.07 |  |

\* denotes $p < .001$ ; \*\* denotes $p < .000$

### Bivariate Results

In bivariate analyses (also in Table 1), those having experienced housing instability were more significantly more likely to have also experienced trauma in the form of former peer victimization and reported significantly less social support. Those who reported housing instability were significantly more likely to report present day unemployment, less likely to have a college degree or health insurance, were more likely to have histories of sex work, had significantly higher levels of peer victimization scores, and report more behavioral risk factors for STI as well as a prior STI in their lifetimes. Within the youth experiencing housing instability sample, there were significant gender differences in former peer victimization scores, with trans male MSM reporting higher levels of peer victimization (M = 19.00, SD = 5.74) than participants identifying as male (M = 14.03, SD = 6.77), trans female (18.22, SD = 3.70), or something else (M = 17.33, SD = 5.06).

Similarly, trans male participants reported higher levels of behavioral risk factors for sexually transmitted infections (M = .50, SD = .25) than those identifying as male (M = .22, SD = .15), trans female (M = .29, SD = .16), or something else (M = .23, SD = .16). Trans male participants also reported lower levels of IPV (M = 16.57, SD = 3.69) than participants identifying as male (20.40, SD = 3.76), trans female (M = 19.09, SD = 3.59), or something else (M = 21.05, SD = 3.92). There were no other significant demographic differences. Mean differences and significance levels of scaled items for youth experiencing housing instability versus those who did not report housing instability, are included in Table 2.

**Table 2.** Descriptive and Bivariate Associations with Housing Instability.

| Item (range) | Experienced housing instability<br>*m (sd)* | Stably housed<br>*m (sd)* | *T (df)* | *p* |
| --- | --- | --- | --- | --- |
| Resilience (0-50) | 28.39 (6.48) | 29.06 (5.91) | 1.93 (1592) | .05 |
| Former peer victimization (0-24) | 14.41 (6.69) | 10.17 (6.36) | -11.06 (1510) | .000 |
| GLBT resources (0-28) | 3.74 (4.2) | 3.11 (3.35) | -3.02 (1546) | .003 |
| Intimate partner violence (0-12) | 20.33 (3.79) | 22.47 (2.53) | 12.55 (1531) | <.001 |
| Behavioral risk factors for sexually transmitted infection (0-6) | .23 (.16) | .18 (.12) | -5.95 (1531) | .000 |
| Social support (1-72) | 46.44 (13.98) | 51.75 (12.84) | 6.88 (1530) | .000 |

\*\*for all scales, higher numbers indicate higher levels of the variable

### Multivariable Analysis

The logistic regression model was statistically significant, χ2 (11, N = 1462) = 270.10, *p* = .001, indicating that the model explained 25% (Nagelkerke R^2^) of the variance in housing instability and correctly classified 79.5% of the cases. As shown in Table 3, seven variables made a statistically significant contribution in the model at the .05 significance level (former peer victimization, gender, age, sex work, IPV, social support, and health insurance status). The strongest predictor of housing instability in our sample reporting sex work in the last three months recorded an odds ratio of 2.76, indicating that young people who reported participating in sex work in the last three months were almost three times as likely to report housing instability. Similarly, behavioral risk factors for sexually transmitted infection (i.e., condomless sex and injectable drug use) recorded an odds ratio of 2.57, but with a significance level of *p* = .07. This indicated that individuals with higher levels of behavioral risk for sexually transmitted infection were more than 2.5 times more likely to report housing instability.

**Table 3.** Multivariate associations with youth housing instability.

|  | Adjusted Odds Ratio (AOR) | AOR 95% CI |  | Scale range, if applicable |
| --- | --- | --- | --- | --- |
| Resilience | 1.00 | .98 | 1.03 | 0-50 |
| Former peer victimization | **1.07** | 1.05 | 1.10 | 0-24 |
| GLBT resources | 1.00 | .96 | 1.04 | 0-28 |
| Behavioral risk factors for STI | **2.57** | .922 | 7.18 | 0-6 |
| IPV scale | **.89** | .85 | .93 | 0-12 |
| Social support | **.98** | .97 | .99 | 1-72 |
| Sex work (1 = yes) | **2.76** | 2.04 | 3.74 |  |
| Health insurance (1 = yes) | **1.63** | 1.21 | 2.21 |  |
| STI lifetime (1 = yes) | **1.21** | .82 | 1.78 |  |
| Age in years | **1.13** | 1.05 | 1.22 |  |
| Cisgender male (1 = yes) | **.34** | .18 | .63 |  |

## Discussion

This study was one of the first to investigate STI risk and infection in the context of housing instability in a large U.S. national sample of young sexual and gender minority (SGM) men having sex with men. We found that engaging in sex work and behavioral risk factors for STI were two of the strongest variables associated with housing instability in our sample. Trauma in the form of former peer victimization was also a significant co-variate in our model, particularly for trans-identified men and women. Not surprisingly, those who experienced housing instability reported less social support than those who did not report housing insecurity. And finally, IPV was also a significant co-variate in our model, though not in the hypothesized direction. MSM youth who experienced housing instability were less likely to report history of IPV. This may be an issue specific to MSM youth who experience housing instability and warrants further exploration. Young MSM might be staying in relationships with violence to avoid housing instability and related additional stigmas.

As hypothesized, our findings that SGM who had sex with men and experienced housing instability were more likely to engage in behaviors that increased vulnerability for STIs, including sex work, was consistent with existing literature on STI risk and the general population of youth experiencing housing instability, including those who identify LGBTQ+^6,7,11^. MSM, including transgender men and women, who experience housing instability, may be more likely to engage in sex work or exchange-sex for basic needs such as food or shelter. Addressing structural barriers to housing and other basic needs, like food insecurity, has the potential to reduce vulnerability to STIs, and merits examining.

Our findings were also consistent with the literature around youth experiencing housing instability and trauma which indicate trauma is highly associated with homelessness and housing instability^5,6,15^. However, former peer victimization is a new angle that has not been studied as extensively in this population. This could be related to intersectionality and sexual and gender minority identification because youth who identify as SGM typically experience more bullying and victimization from peers and others^11,22^. Participating in sex work, a risk factor for STI, is also highly correlated with trauma and housing instability among youth in the literature and our findings support this^4,14^. Youth who experience housing instability and particularly SGM youth experiencing housing instability who have sex with men are a marginalized population with multiple intersecting stigmas, including housing related stigma^24,25^. They may be more likely to be placed in situations that lead them to participate in sex work because of housing problems and related structural inequities that lead to housing instability and homelessness.

Related to peer victimization, we found that young SGM having sex with men who did not report housing instability indicated higher levels of social support. This was consistent with the literature, which indicates that social support is a protective factor for youth, including young people experiencing housing instability^10^. Young MSM with adequate social support, including peers and adults might have more resources and be less likely to experience housing insecurity or feel pressured financially to engage in sex work or sex bartering for basic needs.

## Implications

Our findings indicated that having experienced housing instability was associated with sexually transmitted infections (STIs) and related behavioral risk factors in a large U.S. national cohort of young sexual and gender minority (SGM) men having sex with men. Providing adequate social support is also essential, as young SGM who have sex with men are vulnerable due to multiple intersecting marginalized identities and those who also experience homelessness are at an even higher risk. We found that SGM who had sex with men who experienced housing instability were less likely to have social support. Mentoring and support from adult role models, including case managers who have adequate time to spend cultivating a relationship will be protective of future success for youth experiencing housing instability. It may be helpful to consider matching young SGM who have sex with men with mentors who are also sexual or gender minorities to help address past and current trauma and assist with accessing needed resources.

It is important to educate physicians and other interdisciplinary health care professionals about these risk and protective factors. Developing and implementing training programs that target interprofessional groups across medical settings and within legal/justice and education settings is also a necessity. Clinicians can provide support groups for those involved in sex work to address sexual health safety and social determinants of health to mitigate for those who engage in sex work. For example, assisting with finding housing and financial assistance and food insecurity. Street outreach efforts should include finding youth experiencing housing instability who are doing sex work and offering services where they are. The justice system should implement programming that targets youth experiencing housing instability or homelessness who enter the system, knowing their vulnerabilities and work toward supporting them rather than further criminalizing them. Monies spent on the justice system and policing could be re-routed to support for youth experiencing housing instability and longer term programs that offer adult guidance and mentoring that includes concrete support for housing and food, training for jobs and school. Programs must include funds for staff to help young SGM men having sex with men experiencing housing instability develop coping skills to address trauma that is often ongoing due to their life circumstances while helping them to obtain and maintain safe independent housing with supportive services and a financial safety net. Trauma and PTSD diagnoses will follow people for years and requires long term care and help while in recovery.

## Limitations

Although this was a large U.S. national sample, it was limited to SGM men who were having sex with men, leaving out youth experiencing homelessness who identify as cisgender female and heterosexual males. At least half of the population of youth who experience housing instability, and in some cases more depending on location, identify as LGBTQ+ so the large sample used in this study was meaningful despite its limitations. Also, the data used for this analysis were cross sectional, limiting the ability to make causal inferences. Finally, the data used in this analysis were self-report and does not include objective measures of observation.

## Conclusion and Future Directions

There are opportunities for further research to learn more about larger samples of youth experiencing housing instability who identify as female and heterosexual male. It is essential to understand the largest co-variates of housing insecurity, including STI, among youth across the country and more globally in order to solve this important public health issue. We will not solve large public health issues including the spread of sexually transmitted infections, unless we address social determinants of health, such as housing insecurity. We should conduct further research to learn more about STI in particular among this group, and to learn more about why young people experiencing housing instability more broadly are not engaging in preventive measures. Future research also should focus on developing interventions to improve access to housing and other basic needs for these vulnerable populations. Interventions that reduce stigma around sexual identity and STIs are important for the general public and also to address peer-related bullying. Our findings point to this form of victimization as a significant issue to young SGM men having sex with men experiencing housing instability. Finally, future interventions must address the structural inequities that are driving the housing shortages and lack of resources for young people who experience housing instability or homelessness ^26^. Finding remedies to address health equity and financial insecurity are paramount in communities that have been marginalized because of racial and ethnic identification, socioeconomic status, or other marginalized identities.

In conclusion, addressing factors associated with housing insecurity among one of the most vulnerable groups of adolescents, youth experiencing homelessness and housing insecurity, is crucial to public health and to resolving social determinants of health. SGM men who have sex with men are particularly vulnerable as they experience intersecting stigmas as youth experiencing housing instability, SGM, and often times marginalized race or ethnicity. Using a large U.S. national sample, we have identified several prominent problems that should be imminently addressed in this group, including STI treatment, support for youth who engage in sex work, peer victimization and bullying, and behavioral issues associated with sexually transmitted infection (i.e., injectable drug use, condomless sex). Additionally, improving social support from peers and adult role models is important. Finally, we should further investigate IPV and examine why young SGM men who have sex with men who were stably housed were more likely to experience IPV.

## Data Availability

All data produced in the present study are available upon reasonable request to the authors.

## Acknowledgements

Special thanks to additional members of the T5K study team: Matthew Stief, Drew Westmoreland, Alexa B. D’Angelo, Chloe Mirzayi, Michelle Dearolf, Pedro B. Carneiro, David Pantalone, Adam W. Carrico, Sarit A. Golub, Sabina Hirshfield, Donald R. Hoover, Denis Nash, Gregorio Millett, & Sarah Kulkarni. Thank you to the program staff at NIH: Gerald Sharp, Sonia Lee, Lori Zimand, and Michael Stirratt. And thank you to the members of our Scientific Advisory Board: Michael Camacho, Demetre Daskalakis, Claude Mellins, and Milo Santos. While the NIH financially supported this research, the content is the responsibility of the authors and does not necessarily reflect official views of the NIH.

## Notes

**Funding:** *Together 5,000* (T5K) was funded by the National Institutes of Health (UH3 AI 133675 - PI Grov). Other forms of support include the CUNY Institute for Implementation Science in Population Health, the Einstein, Rockefeller, CUNY Center for AIDS Research (ERC CFAR, P30 AI124414). D.A.W. was supported, in part, by a career development award (K01 AA 029047).

There are no potential conflicts of interest to disclose, real or perceived for any of the authors.

### Competing Interest Statement

The authors have declared no competing interest.

### Funding Statement

Together 5,000 (T5K) was funded by the National Institutes of Health (UH3 AI 133675 - PI Grov). Other forms of support include the CUNY Institute for Implementation Science in Population Health, the Einstein, Rockefeller, CUNY Center for AIDS Research (ERC CFAR, P30 AI124414). D.A.W. was supported, in part, by a career development award (K01 AA 029047).

### Author Declarations

The Institutional Review Board at the City University of New York gave ethical approval for this work.

## References

1. Morton MH, Dworsky A, Matjasko JL, et al. Prevalence and correlates of youth homelessness in the United States. J Adolesc Health. 2018;62(1):14–21.

2. New York City Youth Count.; 2018.

3. Hatchimonji, D. Flatley, C. Treglia, D. Cutuli, J.J. High School Students Experiencing Homelessness: Findings from the 2019 Youth Risk Behavior Surveillance System (YRBSS). Published online 2021. https://files.eric.ed.gov/fulltext/ED616088.pdf

4. Srivastava A, Rusow JA, Holguin M, et al. Exchange and Survival Sex, Dating Apps, Gender Identity, and Sexual Orientation Among Homeless Youth in Los Angeles. J Prim Prev. 10/2019;40(5):561–568.

5. Drescher CF, Griffin JA, Casanova T, et al. Associations of physical and sexual violence victimisation, homelessness, and perceptions of safety with suicidality in a community sample of transgender individuals. Psychology & Sexuality. Published online November 24, 2019:1–12.

6. Eastwood EA, Nace AJ, Hirshfield S, Birnbaum JM. Young Transgender Women of Color: Homelessness, Poverty, Childhood Sexual Abuse and Implications for HIV Care. AIDS Behav. Published online December 21, 2019. doi:10.1007/s10461-019-02753-9

7. Petry L, Hill C, Milburn N, Rice E. Who Is Couch-Surfing and Who Is on the Streets? Disparities Among Racial and Sexual Minority Youth in Experiences of Homelessness. J Adolesc Health. 2022;70(5):743–750.

8. Middleton J, Edwards E, Roe-Sepowitz D, Inman E, Frey LM, Gattis MN. Adverse childhood experiences (ACEs) and homelessness: A critical examination of the association between specific ACEs and sex trafficking among homeless youth in kentuckiana. J Hum Traffick. Published online February 10, 2022:1–14.

9. Côté PB. Sexual Health Services for Homeless Youth: A Qualitative Analysis of their Experiences. J Soc Serv Res. 2019;45(3):429–443.

10. Yoshioka-Maxwell A, Rice E. Exploring the Relationship Between Foster Care Experiences and HIV Risk Behaviors Among a Sample of Homeless Former Foster Youth. AIDS Behav. 3/2019;23(3):792–801.

11. Heerde JA, Hemphill SA. Sexual Risk Behaviors, Sexual Offenses, and Sexual Victimization Among Homeless Youth: A Systematic Review of Associations With Substance Use. Trauma Violence Abuse. 12/2016;17(5):468–489.

12. Guilamo-Ramos V, Thimm-Kaiser M, Benzekri A, Futterman D. Youth at risk of HIV: the overlooked US HIV prevention crisis. Lancet HIV. 2019;6(5):e275–e278.

13. Cdc. HIV surveillance report: Diagnoses of HIV infection in the United States and dependent areas. Published online 2016.

14. Halverson M, Hatchimonji DR, Treglia D, Flatley CA, Herbers JE, Cutuli JJ. Risky sexual behavior and STI testing among teens experiencing homelessness. Child Youth Serv Rev. 2022;139(106538):106538.

15. Wong CF, Clark LF, Marlotte L. The Impact of Specific and Complex Trauma on the Mental Health of Homeless Youth. J Interpers Violence. 03/2016;31(5):831–854.

16. Grov C, Westmoreland DA, Carneiro PB, et al. Recruiting vulnerable populations to participate in HIV prevention research: findings from the Together 5000 cohort study. Ann Epidemiol. 2019;35:4–11.

17. Nash D, Stief M, MacCrate C, et al. A Web-based study of HIV prevention in the era of pre-exposure prophylaxis among vulnerable HIV-negative gay and bisexual men, transmen, and transwomen who have sex with men: Protocol for an observational cohort study. JMIR Res Protoc. 2019;8(9):e13715.

18. Grov C, Westmoreland DA, Carrico AW, Nash D. Are we on the precipice of a new epidemic? Risk for hepatitis C among HIV-negative men-, trans women-, and trans men-who have sex with men in the United States. AIDS Care. 2020;32(sup2):74–82.

19. Connor KM, Davidson JRT. Development of a new resilience scale: the Connor-Davidson Resilience Scale (CD-RISC). Depress Anxiety. 2003;18(2):76–82.

20. Zimet GD, Powell SS, Farley GK, Werkman S, Berkoff KA. Psychometric characteristics of the Multidimensional Scale of Perceived Social Support. J Pers Assess. 1990;55(3-4):610–617.

21. Straus MA, Hamby SL, BONEY-McCOY S, Sugarman DB. The Revised Conflict Tactics Scales (CTS2): Development and Preliminary Psychometric Data. J Fam Issues. 1996;17(3):283–316.

22. Greenwood GL, Relf MV, Huang B, Pollack LM, Canchola JA, Catania JA. Battering victimization among a probability-based sample of men who have sex with men. Am J Public Health. 2002;92(12):1964–1969.

23. Joseph S, Stockton H. The multidimensional peer victimization scale: A systematic review. Aggress Violent Behav. 2018;42:96–114.

24. Sisselman-Borgia A, Budescu M, Torino G. The association of Racial and homelessness microaggressions and physical and mental health in a sample of homeless youth. Urban Soc Work. 2018;2(2):139–158.

25. Sisselman-Borgia A, Menard L, Budescu M, Torino G. Differences in Discrimination Experiences Among Homeless and Nonhomeless Youth. Urban Social Work. 2022;6(1):69–83.

26. Olivet J, Wilkey C, Richard M, et al. Racial Inequity and Homelessness: Findings from the SPARC Study. Ann Am Acad Pol Soc Sci. 2021;693(1):82–100.

